# Tension between timeliness and completeness of data in the initiation of cancer treatment: A qualitative study of oncology practice workflows and enduring health IT challenges

**DOI:** 10.1101/2025.05.19.25324967

**Authors:** Lipika Samal, Michael Anne Kyle, John L. Kilgallon, Kristen McNiff Landrum, Atul A. Gawande, Joseph O. Jacobson, Michael J. Hassett

**Author notes:** Corresponding author: Lipika Samal, MD, MPH, Division of General Internal Medicine and Primary Care, Brigham and Women’s Hospital, 1620 Tremont St., Suite OBC-03-02V Boston, MA 02120. **Ethics:** This study was reviewed and approved by the Partners HealthCare Human Research Committee.

## Abstract

**Introduction:** Diagnostic evaluation and treatment planning for newly diagnosed cancer requires a coordinated effort across multiple specialties. Delays in treatment initiation are common, leading to unnecessary anxiety and decreased survival. Given that timely treatment initiation is pivotal to providing high quality cancer care, we sought to characterize patient intake, workflows, and the role of health information technology (HIT) in a varied group of oncology practices nationwide.

**Methods:** Interviews with oncologists were performed between March and September 2016, with follow-ups conducted between October and December 2021. Thematic analysis was used to assign codes to key elements of the transcripts, group these codes into conceptually distinct and clinically meaningful categories, and identify major cross-cutting themes.

**Results:** Nine oncologists participated in an initial interview (one surgical, two radiation, six medical oncology). Four oncologists participated in a follow-up interview (one radiation, three medical oncology). In both time periods there was tremendous variation in staff roles and communication processes; some oncology practices obtained diagnostic studies before the first oncology consult visit, whereas others waited until after the initial consult visit to begin the diagnostic evaluation. Variability and tension were noted to arise from deficiencies in HIT, such as lack of interoperability, impaired speed and quality of data collection, cumbersome user interfaces, and variety of data types in oncology care. Oncologists reported only modest improvements in HIT between 2016 and 2021.

**Conclusion:** Assembling data to make a new cancer diagnosis and treatment plan is complex and time-intensive. HIT interoperability remains a quasi-manual process, contributing to preventable treatment delays. Federal policy supporting interoperability provides an opportunity to develop HIT that supports care coordination and patient-centered care, but effective implementation of such tools will be challenging within current workflows.

## Introduction

Delays in initiating cancer treatment have been associated with higher mortality for lung cancer and colorectal cancer patients and worse outcomes for breast cancer patients.^1-3^ In some settings, the magnitude of the increase in mortality nullifies the survival benefits attributed to modern chemotherapy.^4^ Across all three cancer treatment modalities of surgery, systemic treatment, and radiotherapy, a delay of just four weeks has been shown to carry a significantly increased risk of death, and every additional month delay of surgery is associated with a 6-8% increase in mortality.^5^

Evidence reveals wide variability in the length of time from a new cancer diagnosis to initiation of treatment; in one study of lung cancer treatment, the time from the first oncology consult visit to treatment ranged from 1 to 1,687 days.^6^ Moreover, the magnitude of delay is increasing over time, perhaps due to the growing complexity of cancer care.^7,8^ Despite the importance of timely access to cancer care, few studies have examined the process in order to determine which factors contribute to delays, and this has been identified as a priority area for research.^9^ In particular, the diagnostic evaluation and treatment planning period is an understudied stage of care.

This study explores how oncologists in diverse settings intake new patients and the practice workflows and health information technology (HIT) that support them. HIT tools are evolving to include oncology-specific features of electronic health records (EHRs), oncology interoperability standards, radiation oncology systems that function as EHRs, second opinion platforms, video tumor board software, and other tools.^10,11^ Considering that the diagnostic evaluation and treatment planning period is pivotal to providing high quality cancer care, we sought to characterize new patient intake, clinical workflows, and the role of HIT for tasks such as: 1) review of certain types of clinical data (e.g., radiology, pathology, lab values, medications, progress notes from primary care and other specialties), 2) ordering and tracking of diagnostic test results, 3) communication, and 4) clinical documentation, with the goal of informing the development of HIT tools that take advantage of new national efforts to improve interoperability.

## Methods

We interviewed oncologists to understand their workflow during the diagnostic evaluation and treatment planning period of care. We recruited medical, surgical, and radiation solid-tumor oncologists from a variety of settings nationally, including sites receiving referrals directly from primary care and sites providing second opinions. We focused on solid tumors because these cases frequently require communication and coordination across multiple oncologic specialties. We used snowball sampling to identify participants using our own contacts and contacts recommended by participants. Initial interviews were conducted March-September 2016 and follow-up interviews were conducted October-December 2021.

We developed a semi-structured interview guide based on a literature review, our own clinical experience, meetings with Oncology HIT vendors and advice from a Technical Expert Panel. Two investigators conducted every interview by telephone. All interviews were audio-taped and transcribed verbatim. The study was deemed exempt by the Partners HealthCare Human Research Committee.

We used thematic analysis to assign codes to key elements of the transcripts, to group these codes into conceptually distinct and clinically meaningful categories, and to identify major cross-cutting themes.^12^ We used an open coding process and constant comparative method of analysis.^13^ Data were iteratively coded. Two investigators (LS, MH, or JOJ) reviewed each transcript and developed a codebook. Each transcript was re-coded by one or two investigators (LS, MH, MAK) to review and refine conceptual themes.

## Results

Nine oncologists participated in an initial interview (one surgical, two radiation, six medical oncology). Four oncologists participated in a follow-up interview (one radiation, three medical oncology), of which two oncologists were recruited from additional sites (Appendix).

### Overview: New Patient Treatment Initiation Processes Varied Extensively Across Sites

We divided the diagnostic evaluation and treatment planning period into four phases with emphasis on the tasks that occur before the initial consultation visit with an oncologist: 1) Initial Triage, from initial referral to the act of scheduling the first consult appointment with a cancer specialist; 2) Pre-consult, from scheduling of the first consult appointment to the first consult visit; 3) Consult, the initial consultation visit; and 4) Post-consult, from the first consult visit to initiation of treatment (Figure 1).

**Figure 1.**
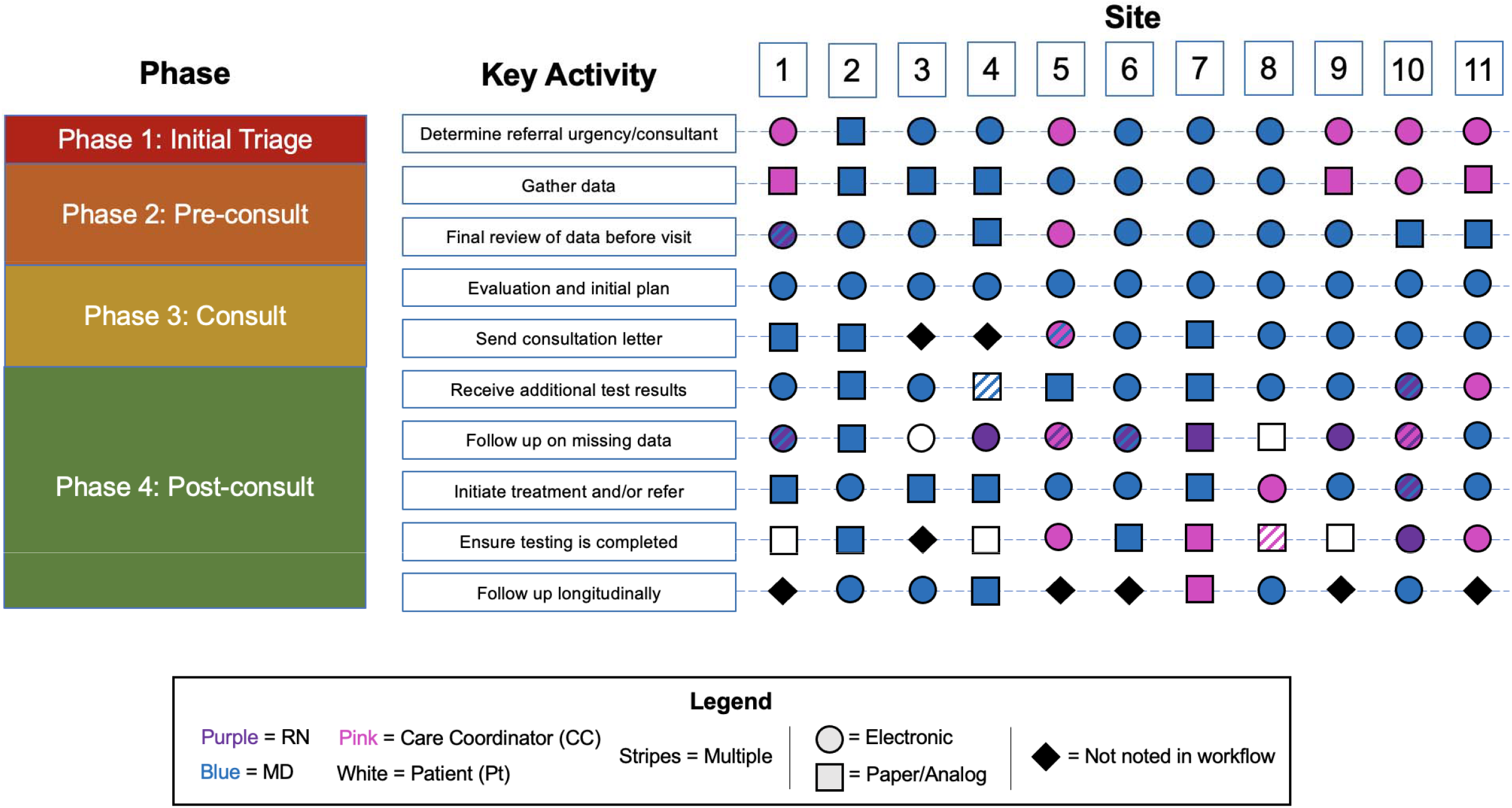
Variation in use of health information technology for key activities and cancer care delivery practices across eleven sites. We identified 10 key tasks performed during the diagnostic evaluation and treatment planning period; the role of the individual responsible for the task (e.g., MD, RN, care coordinator) and whether the task was performed electronically (within the EHR) or analog (communication or documentation outside of the EHR including scanned, non-searchable PDFs uploaded to an electronic record) are indicated above. Phases include: 1) Initial Triage, from initial referral to the act of scheduling the first consult appointment with a cancer specialist; 2) Pre-consult, from scheduling of the first consult appointment to the first consult visit; 3) Consult, the initial consultation visit; and 4) Post-consult, from the first consult visit to initiation of treatment.

We identified 10 key tasks performed during the treatment initiation. We assigned each task to a phase of care initiation, and we indicated the role of the individual responsible for the task (e.g., MD, RN, care coordinator) and, where applicable, whether the task was performed electronically (within the EHR) or analog (communication or documentation outside of the EHR including scanned, non-searchable PDFs uploaded to an electronic record). The interviews revealed tremendous heterogeneity in the roles of physicians and clinic staff, as well the use of digital and analog sources of clinical data. The qualitative data provided insight into the efficiencies and inefficiencies in each of the four phases (Table 1).

**Table 1.**
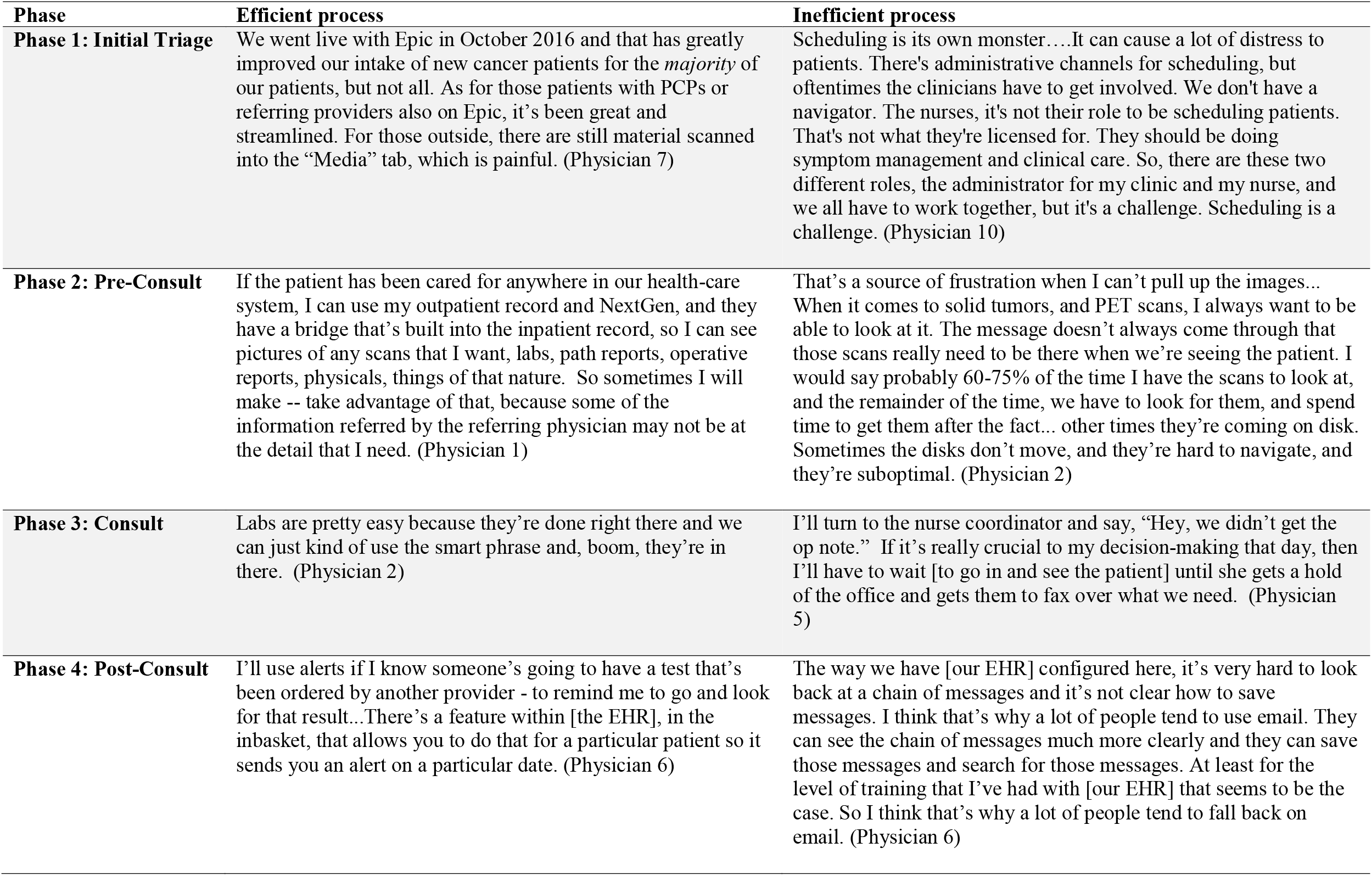
Efficiencies and Inefficiencies of Clinical Information Systems and Workflows in Each Phase.

| Phase | Efficient process | Inefficient process |
| --- | --- | --- |
| **Phase 1: Initial Triage** | We went live with Epic in October 2016 and that has greatly improved our intake of new cancer patients for the *majority* of our patients, but not all. As for those patients with PCPs or referring providers also on Epic, it's been great and streamlined. For those outside, there are still material scanned into the "Media" tab, which is painful. (Physician 7) | Scheduling is its own monster....It can cause a lot of distress to patients. There's administrative channels for scheduling, but oftentimes the clinicians have to get involved. We don't have a navigator. The nurses, it's not their role to be scheduling patients. That's not what they're licensed for. They should be doing symptom management and clinical care. So, there are these two different roles, the administrator for my clinic and my nurse, and we all have to work together, but it's a challenge. Scheduling is a challenge. (Physician 10) |
| **Phase 2: Pre-Consult** | If the patient has been cared for anywhere in our health-care system, I can use my outpatient record and NextGen, and they have a bridge that's built into the inpatient record, so I can see pictures of any scans that I want, labs, path reports, operative reports, physicals, things of that nature. So sometimes I will make -- take advantage of that, because some of the information referred by the referring physician may not be at the detail that I need. (Physician 1) | That's a source of frustration when I can't pull up the images... When it comes to solid tumors, and PET scans, I always want to be able to look at it. The message doesn't always come through that those scans really need to be there when we're seeing the patient. I would say probably 60-75% of the time I have the scans to look at, and the remainder of the time, we have to look for them, and spend time to get them after the fact... other times they're coming on disk. Sometimes the disks don't move, and they're hard to navigate, and they're suboptimal. (Physician 2) |
| **Phase 3: Consult** | Labs are pretty easy because they're done right there and we can just kind of use the smart phrase and, boom, they're in there. (Physician 2) | I'll turn to the nurse coordinator and say, "Hey, we didn't get the op note." If it's really crucial to my decision-making that day, then I'll have to wait [to go in and see the patient] until she gets a hold of the office and gets them to fax over what we need. (Physician 5) |
| **Phase 4: Post-Consult** | I'll use alerts if I know someone's going to have a test that's been ordered by another provider - to remind me to go and look for that result...There's a feature within [the EHR], in the inbasket, that allows you to do that for a particular patient so it sends you an alert on a particular date. (Physician 6) | The way we have [our EHR] configured here, it's very hard to look back at a chain of messages and it's not clear how to save messages. I think that's why a lot of people tend to use email. They can see the chain of messages much more clearly and they can save those messages and search for those messages. At least for the level of training that I've had with [our EHR] that seems to be the case. So I think that's why a lot of people tend to fall back on email. (Physician 6) |

**Table 2.** Sample Characteristics.

| **Site** | **Oncology type** | **Gender** | **Years in practice** | **Practice Setting** | **Primary Catchment Area** | **Region** |
| --- | --- | --- | --- | --- | --- | --- |
| **1** | Medical (General) | M | More than 30 | Independent community practice | Urban, small metro | Midwest |
| **2** | Medical (General) | F | 10 to 19 | Health system-affiliated community practice | Suburban | Northeast |
| **3** | Radiation | M | 20 to 29 | Health system-affiliated community practice | Suburban | Northeast |
| **4** | Surgical | F | 10 to 19 | NCI Comprehensive Cancer Center | Rural | Northeast |
| **5** | Medical (General) | M | 20 to 29 | NCI Comprehensive Cancer Center | Urban, large metro | Midwest |
| **6** | Radiation | M | 20 to 29 | NCI Comprehensive Cancer Center | Suburban | Midwest |
| **7** | Medical (General) | M | 10 to 19 | Health system-affiliated community practice | Rural | West |
| **8** | Medical (General) | F | 30 to 39 | Integrated Delivery Network | Urban, large metro | West |
| **9** | Medical (General) | M | 10 to 19 | Health system-affiliated community practice | Rural | West |
| **10** | Medical (Thoracic) | M | Less than 10 | NCI Comprehensive Cancer Center | Urban, large metro | Northeast |
| **11** | Medical (Breast) | M | 10 to 19 | NCI Comprehensive Cancer Center | Urban, large metro | Northeast |

### Phase 1: Initial Triage

During initial triage, the urgency of the incoming consultation request was established. Most practices received the initial consult request electronically, where it was processed either by an oncologist (in smaller practices) or trained intake coordinators (in larger organizations).

Patients presenting with acute, fast-growing cancers were triaged quickly; it was clear they needed to initiate oncology care as soon as possible (within days). More commonly, patients presented with cancers that required prompt but not urgent attention (within 2-3 weeks). Oncologists then needed to decide how to balance speed and competeness of data. Oncologists noted having preparation time before the initial consultation could be valuable, as it provided an opportunity to assemble clinical data prior to the initial consultation, during which they knew patients would be eager for a definitive diagnosis and treatment options. However, oncologists were attuned to the distress imposed by a new cancer diagnosis and desired to form a relationship with the patient as soon as possible.

> *It is a balance between getting the patient in expeditiously versus making sure that every single last piece of data are obtained before the patient comes in. Oftentimes, I will see the patient without [complete] data, to assuage concerns with waiting around and how long it takes to see ‘somebody who can help them,’ (Physician 4)*

All oncologists, regardless of specialty, described negotiating the tension between having a rapid, incomplete initial visit versus a slightly delayed, more complete initial visit. Oncologists’ prioritization of speed versus complete information shaped their approach to subsequent phases of intake. While clinical factors mattered most, oncologists at smaller practices serving less urban populations emphasized completeness, whereas those at major urban referral centers emphasized speed.

### Phase 2: Pre-consult

Gathering data was the central pre-consult phase activity for all practices, though they went about data collection in a variety of ways. The workflow in some sites included clinic staff actively compiling a complete set of diagnostic information from multiple clinical settings. At these sites the first consult appointment was usually scheduled after pending medical record requests were completed. The efficiency and effectiveness of the process depended on the medical knowledge base of staff and the opportunity for staff to consult with an oncologist about what information would be needed for diagnosis (e.g., results of key genetic tests). The type of clinic staff involved in pre-consultation collection and review of clinical data varied greatly across sites, with some utilizing nurses, others patient liaisons, and others relying entirely on the treating physician. For example, one oncologist had a team of two staff, a nurse navigator and administrative assistant, who worked together to gather records into a packet of emailed or printed records before presenting this information to a physician. Conversely, in another practice it was the oncologist’s responsibility to determine the extent of any prior workups and case urgency before the initial consultation.

The extent of pre-consult data collection depended on the extent to which speed or information was prioritized in scheduling the initial visit, but it also depended heavily on the accessibility of patients’ medical records. Patients within the same health system might have pre-consultation documentation assembled immediately, whereas external patients with records on a compatible EHR (e.g., Epic Care Everywhere) could have at least some information assembled immediately, and patients whose records relied entirely on external records might require considerable follow-up, including manual collection of records by patients themselves.

In addition to assembling existing documentation, oncologists varied in their approach to ordering additional diagnostic testing prior to the initial consult (Appendix B). Two oncologists scheduled diagnostic procedures, such as endoscopic ultrasound or lung biopsy, before the first consult visit. Others waited to meet the patient first.

### Phase 3: Consult

We defined the consult phase, or initial consultation, as the first visit between the new patient and the oncologist. Oncologists approached these evaluations similarly: they met the patient, initiated a relationship, synthesized information obtained pre-consult with information obtained in the visit, and identified outstanding gaps in information. During this meeting, the oncologist completed their initial evaluation and plan, followed by a consult note. They consistently described taking great care in preparing the initial consult note, using it as a template for their own treatment plan as well as a key mode of communication with other colleagues on the medical team. In all cases, the treating oncologist conducted the initial evaluation and plan in-person. The consultation note was always written by the treating oncologist, with some variation in mode of dissemination to other members of the patient’s team. The ability to create a treatment plan was dependent on the completeness of the evaluation at the time of the visit. If comprehensive diagnostic information was available and assembled prior to the visit, the plan could proceed to treatment. If the visit took place without complete information, the post-consult phase focused on filling outstanding gaps in data, including continuing to gather records, completing more diagnostic testing, obtaining input from oncologist colleagues. A key insight was that practices with more extensive support systems for the initial triage and pre-consult phases were able to move patients into treatment immediately after the initial consultation visit, whereas practices with less extensive support prior to the initial consultation required more time to initiate treatment.

### Phase 4: Post-Consult; Treatment Planning and Initiation

The post-consult phase had the most variation in terms of tasks and workflows. In some practices, HIT improved timeliness of treatment initiation. For example, if a patient proceeded directly to treatment, the oncologist would activate the relevant order set in the EHR, which would in turn populate the workflows of the other parties involved in executing the treatment plan. In other practices with less extensive support systems prior to the initial consultation visit, the hallmark of the tasks included in the post-consult phase included receiving outstanding test results and following up on missing data.

While oncologists played a role in identifying missing information, other practice staff, such as nurses or navigators, were responsible for tracking down follow-up data. Similarly, scheduling and follow-up of new testing was generally completed by practice staff. If a patient required consultations from other oncologic specialties, the oncologist would often contact colleagues directly, though sometimes patient navigators would be delegated to circulate information.

For oncologists, the post-consult tasks represented the greatest opportunity to delegate work. However, the variation that was a hallmark of this phase created its own challenges. For instance, communicating with other physicians and team-members to develop and initiate the treatment plan was the key task of oncologists during the post-consult phase. Oncologists described the many possible channels for peer-to-peer communication as a source of frustration. Oncologists could send and receive communications via work email, EHR email, secure texting, phone, letter, fax, or in person. The variability of information sources was a source of stress.

> *It is a little frustrating to me that we don’t have a standardized way of communicating. I’d prefer that we stop using email and use the inbasket. Or we stop using the inbasket and we use email. It’s just another thing to check. (Physician 6)*

The challenge of multiple channels did not improve over time. The above quote from an oncologist in 2016 mirrored the below quote from a different oncologist in 2021.

> *They’ve tried different systems here. We have something called Voalte, Microsoft Teams—but you’ve all got to be on it…*.*There’s a lot of ways you get communicated to, but the only one that’s widespread is email. It starts to create a lot of cognitive load to be communicated to through so many different platforms because then you’re like, what am I monitoring? (Physician 10)*

## HIT Contributes to Tension Between Complete Diagnostic Workup and Timely Treatment Initiation

While some of the tension between speed and information arose from genuine clinical issues, such as determining the need for and completing further testing, much of this tension arises from deficiencies in HIT. The issues were manifold: lack of interoperability (“between EHR”) impaired speed and quality of data collection, the user interface of EHRs (“within EHR”) was frustrating, and the variety of data types used in oncology care created unique challenges. Oncologists reported only modest improvements in HIT between 2016-2021.

> *When you got in touch with me again, I was like, wow, five years. And, I was disappointed at how … little had changed, which is very discouraging to me because I think of machine learning and [artificial intelligence] and this is what we’re still dealing with. (Physician 6)*

Both between- and within-EHR interoperability affected timely access to data. Between 2016-2021, oncologists reported that interoperability challenges were somewhat ameliorated by the growing market dominance of certain vendors (e.g., Epic). Yet, despite the widespread adoption and acceptance of EHRs, “paper” problems, such as the difficulty of reviewing scanned documents, were mentioned multiple times. An EHR misuse problem that posed similar data-sifting challenges as unsearchable PDFs was the proliferation of excessive copying and pasting in the electronic chart (“note bloat”).

In addition to challenges around information synthesis in the EHR, oncologists described navigating within the EHR as cumbersome. Many had developed workarounds—often paper-based—to deal with functional shortcomings in their electronic records. Along with printing out documentation, multiple oncologists used “shadow charting” to track critical information outside of the EHR. As one described it, “*I do have a separate checking sheet. And I usually just keep it with my patient stuff. It’s like a ringed notepad with checkboxes*.*”* (Physician 4)

Oncologists faced interoperability and quality challenges beyond standard text-based chart documentation. Oncologists frequently required data such as original imaging and pathology that often did not transmit with their associated reports, and/or were low-quality copies that couldn’t be evaluated (Table 1). In addition to classic imaging and pathology, oncologists increasingly rely on specialty data, such as genetic testing. In a 2021 follow-up, one oncologist identified this as an area where they welcome guidance on solutions: “*I’d ask if anyone has a solution for the next generation sequencing reports? The [vendor] reports get buried in the Media tab and right now we just manually transcribe it into our notes or not at all*.*”* (Physician 7)

Radiation oncologists had unique data needs. They used a separate EHR for programming their therapies, and double-charted information there and the primary EHR. The second system introduced its own interoperability challenges, wherein a radiation oncologist treating a patient with past history of radiation would need precise parameters of prior treatments that were not transmitted via sharing of the primary record alone.

## Discussion

This qualitative study explores practicing oncologists’ clinical workflows and use of HIT in diagnostic evaluation and treatment planning for oncology patients over a period of five years. Five-year follow-up data reveals that there has been some progress due to the consolidation of data in one vendor in the ambulatory setting (Epic) and the addition of patient navigators to the care team. However, oncologists reported only modest progress in timely receipt of diagnostic testing over the study interval. Our findings corroborate previous qualitative work documenting the importance of care coordination in cancer care and enduring challenges related to HIT tools and workflow design.^14,15^

Our qualitative findings provide context for results of previous quantitative studies showing increasing delays in time to treatment. The process of compiling information is still primarily manual and prone to error. Though electronic health records are now standard, information exchange frequently involves outreach to obtain hard-copy documents that are scanned into the chart, a labor-intensive process resulting in data that is not truly digital, such as non-searchable PDFs. The lack of true data interoperability continues to represent the main HIT obstacle to safe and efficient care.

There is tension in balancing the goal of having patients seen as quickly as possible by an oncologist as opposed to the goal of complete clinical information to make the patient’s visit as productive as possible. In the present study, oncologists prioritized seeing the patient as quickly as possible but acknowledged the difficult tradeoff. Intake processes were notable for their astounding variety in the number of people involved, their roles, and their use of HIT, or lack thereof. Notably, physicians are deeply involved in parts of the process that could be managed by other care team members or potentially could be automated. Establishing best practices in staffing (team-based care) and workflow standardization/optimization could alleviate burdens on practice staff and physician time especially.^16^ Improved data interoperability could relieve manual workflow burdens and increase timeliness of cancer care initiation by automating adjudication and deduplication of data, particularly from specialized sources like imaging and next-generation sequencing. The National Cancer Institute Informatics Technology for Cancer Research Program has identified these areas as a research priority.^17^

Our study has limitations. Qualitative data are not generalizable. Interviews represent a small sample of cancer specialists and not all initial sample responded to follow-up inquiries; however, we did succeed in recruiting multiple specialties from diverse practices around the country. Finally, while this work emphasized the oncologist perspective on cancer care initiation, there are major implications for other team members and patient experience.^18,19^

Inefficient information exchange burdens patients—often inequitably.^20^ Patients may participate in collecting information tasks and can be stressed by delays. The Centers for Medicare and Medicaid Services has recently issued a proposed rule advancing interoperability which would include a patient-facing view of the status of processes such as prior authorizations—this would give patients insight into the work underway between encounters.^21^

The US healthcare system has not placed enough emphasis on streamlined, error-free healthcare delivery in the ambulatory setting. As the results of this study illustrate, there is potential for an HIT tool that would allow collaboration between oncologic specialties and provide patients a birds’ eye view of the process, similar to HIT tools developed for patients with multiple chronic conditions.^22,23^ However, current HIT tools for multiple chronic conditions have not been proven to be effective and clinicians consider the technology to be disruptive.^20^

## Conclusion

Development of new HIT tools should focus on improving the process of compiling data prior to the initial oncology consult and could potentially resolve the tension between timeliness and completeness of data in the initiation of cancer treatment. Practicing oncologists can support these efforts by advocating for federal health policies that encourage interoperability, as well as leading healthcare delivery system innovation including development of HIT tools.

## Data Availability

All data produced in the present study are available upon reasonable request to the authors

## Acknowledgements

Interview respondents. Ariadne Advisory Board: William Berry, Lisa Hirschhorn, and Natalie Heinrich. Project staff: Joy Gulla, Hillary Teed, Susan Czajak, and Robert Mersereau. Technical Expert Panel: Russell Leftwich, Joan Ash, Debra Patt, Nancy Keating, Shirley Johnson, Gabriela Spear

## Notes

**Funding Statement:** This work was supported by a Spark grant from Ariadne Labs. Dr. Samal has received funding from the National Institutes of Health under R01DK116898 and IBM through a PI- initiated grant for COVID-19 artificial intelligence research, both unrelated to the current work. Dr. Kyle was supported by a National Cancer Institute training grant (5T32CA092203 and K99CA277367). The content is solely the responsibility of the authors and does not necessarily represent the official views of the National Institutes of Health.

### Competing Interest Statement

The authors have declared no competing interest.

### Funding Statement

This work was supported by a Spark grant from Ariadne Labs. Dr. Samal has received funding from the National Institutes of Health under R01DK116898 and IBM through a PI-initiated grant for COVID-19 artificial intelligence research, both unrelated to the current work. Dr. Kyle was supported by a National Cancer Institute training grant (5T32CA092203 and K99CA277367). The content is solely the responsibility of the authors and does not necessarily represent the official views of the National Institutes of Health.

### Author Declarations

This study was reviewed and approved by the Partners HealthCare Human Research Committee

